# Study Design and Rationale for the PAASIM Project, a Matched Cohort Study on Urban Water Supply Improvements and Infant Enteric Pathogen Infection, Gut Microbiome Development, and Health in Mozambique

**DOI:** 10.1101/2022.10.28.22281675

**Authors:** Karen Levy, Joshua V. Garn, Zaida Adriano, Bacelar de Barros, Christine S. Fagnant-Sperati, Sydney Hubbard, Antonio Júnior, João Luís Manuel, Magalhães Mangamela, Sandy McGunegill, Molly K. Miller-Petrie, Jedidiah S. Snyder, Courtney Victor, Lance Waller, Konstantinos T. Konstantinidis, Thomas Clasen, Joe Brown, Rassul Nalá, Matthew C. Freeman

## Abstract

**Introduction:** Despite clear linkages between provision of clean water and improvements in child health, limited information exists about the health impacts of large water infrastructure improvements in low-income settings. Billions of dollars are spent annually to improve urban water supply, and rigorous evaluation of these improvements, especially targeting informal settlements, is critical to guide policy and investment strategies. Objective measures of infection and exposure to pathogens, and measures of gut function, are needed to understand the effectiveness and impact of water supply improvements.

**Methods and analysis:** In the PAASIM study, we examine the impact of water system improvements on acute and chronic health outcomes in children in a low-income urban area of Beira, Mozambique, comprising 62 sub-neighborhoods and ∼26,300 households. This prospective matched cohort study follows 548 mother-child dyads from late pregnancy through 12 months of age. Primary outcomes include measures of enteric pathogen infections, gut microbiome composition, and source drinking water microbiological quality, measured at the child’s 12 month visit. Additional outcomes include diarrhea prevalence, child growth, previous enteric pathogen exposure, child mortality, and various measures of water access and quality. Our analyses will compare a) subjects living in sub-neighborhoods with the improved water to those living in sub-neighborhoods without these improvements; and b) subjects with household water connections on their premises to those without such a connection. This study will provide critical information to understand how to optimize investments for improving child health, filling the information gap about the impact of piped water provision to low-income urban households, using novel gastrointestinal disease outcomes.

**Ethics and dissemination:** The study was approved by the Emory University Institutional Review Board and the National Bio-Ethics Committee for Health in Mozambique. The pre-analysis is published on the Open Science Framework platform (https://osf.io/4rkn6/). Results will be shared with relevant stakeholders locally, and through publications.

**STRENGTHS AND LIMITATIONS OF THE STUDY:**

- This matched cohort study of an urban water supply improvement project will provide critical information about the health impacts of providing piped water and household connections to low-income households.
- We employ rigorous measures of exposure and novel and objective outcome measures, including gut microbiome composition and molecular detection of enteric pathogens.
- The study design allows for examination of both neighborhood and household-level effects of water supply improvements.
- As a natural experiment, we are unable to randomize the intervention, leading to potential residual confounding.
- We are unable to examine the impacts of all aspects of the city-wide water improvement project, due to lack of comparable populations, and instead focus only on the low income neighborhoods.

## 1. INTRODUCTION

Large-scale provision of disinfected, treated drinking water is considered one of the greatest public health achievements of the 20^th^ century[1] and played an important role in improving child health in high-income countries.[2] In low-income countries with high burdens of infectious diseases, inadequate water, sanitation, and hygiene (WASH) conditions are strongly associated with poor child health outcomes, including diarrheal diseases, responsible for >400,000 deaths of children <5 annually,[3,4], and linear growth faltering, underlying 15-17% of mortality of children <5.[5] In Mozambique 27% of stunting is attributed to unimproved water and sanitation.[6]

### 1.1. Robust studies of the health impacts of community water supply are needed

Rapid urbanization is occurring globally, with urban areas expected to account for 96% of the additional 1.4 billion human population by 2030[7] and 68% of the global population expected to live in urban areas by 2050.[8] While sub-Saharan Africa is still predominantly rural, by 2050 the continent is projected to be 56% urban.[9] To cope with urban growth, expanded infrastructure and services in cities and peri-urban areas will be essential.[7] In 2014 alone, over US$4.4 billion was committed to WASH in sub-Saharan Africa, with the majority going to improve water supply.[10] Implementation challenges in lower-income settings—such as intermittent service and pathogen intrusion in the distribution system due to pipe breaks, pressure drops, or illegal connections—limit the potential for engineered systems to provide a continuous supply of treated drinking water directly to homes, in adequate quantities to improve hygiene.[11,12] Given the considerable investment in providing piped services to low-income communities, rigorous evaluation of community-scale water provision is critical, to understand the real-world effectiveness and health impact of such systems in low-income contexts.[10,13,14]

Despite the clear biological link between safe water and child health and development, limited information exists about the health impacts of large water infrastructure improvements in low-income settings. A small number of studies have evaluated upgrades from intermittent to continuous water delivery in urban areas,[15] or localized improvements to water quality at shared water points.[16] Other studies have evaluated sanitation interventions, without examining drinking water or combined water and sanitation interventions.[17] A recent review of interventions to improve water quality globally found no studies evaluating reliable piped-in water supplies delivered to households and specifically called for rigorous research to assess the health impact of reticulated water supply systems.[18] The review concluded that “there is currently insufficient evidence to know if source-based improvements such as protected wells, communal tap stands, or chlorination/filtration of community sources consistently reduce diarrhoea.” A World Health Organization (WHO) review of drinking water and sanitation on diarrhoeal disease in low- and middle-income settings concurs, stating that “evidence from well-conducted intervention studies assessing exclusive use of adequate access and supply of safe water…is still very limited.”[19] One reason for this limited evidence is that community-scale interventions are difficult to study using randomized control trial (RCT) methodology – the gold standard for causal inference. It is often infeasible to randomize intervention groups due to policy, planning, and engineering considerations, and lack of adequate comparison groups. As such, alternative quasi-experimental designs must be applied.[20–22]

The predominance of studies in the WASH sector have focused on household- and compound-level interventions because they lend themselves more readily to RCT methodology. Outside of the few aforementioned studies of community-wide infrastructure improvements, evaluations of the health impact of water quality improvements in low-income settings often focus on household water treatment such as boiling, chlorinating or filtering water, with studies predominantly conducted in rural settings.[18] Results of these trials have been mixed, since household-based approaches have various limitations, including low uptake and inconsistent use,[23,24] post-treatment contamination,[25–28] and a poor record of sustained use.[29,30] Household water treatment interventions do not increase water quantity and availability and typical household WASH interventions are likely insufficient to prevent growth faltering in most cases.[31–34]

It is crucial to assess the impact of community-scale infrastructure improvements,[21] as this is an area that is particularly relevant to inform aid agencies, development banks, and other policy makers.[35] The area of most rapid growth in water access is via piped water supply connections, not household water treatment, and larger infrastructure interventions are also critical to achieving the scale of water supply improvements necessary to make impactful changes.[36]

### 1.2. Objective measures of gut health are needed

A vast majority of WASH studies use as their primary health outcome caregiver-reported diarrhea, primarily because acute diarrheal illness is responsible for ∼10-12% of all deaths in children <5.[37,38] However, diarrhea is an unreliable outcome due to courtesy, social desirability and recall bias,[39] local definitions of diarrhea,[40–44] other self-reporting issues,[39,45–47] and the multiple potential etiologies of diarrhea symptoms.[48] Such biases are especially problematic where interventions cannot be blinded as is mainly the case for water interventions. Shedding of enteropathogens, organisms that cause acute gastrointestinal illness, provides an unambiguous indicator of current infection, and increasingly is being used in the WASH field.[31,49,50] Advances in diagnostic techniques make it feasible to test for a wide variety of enteric pathogens simultaneously.[51,52] It is useful to understand enteric pathogen infections because chronic and repeated enteric pathogen infections in the first two years of life—with or without symptomatic diarrhea—are associated with serious morbidities, including gut impairment, growth shortfalls, and cognitive deficits by ages 7-9 years.[53–59] Such outcomes can have profound impact on health, development and well-being of individuals, communities, and entire countries.[60,61] Host-level gastrointestinal conditions affected by environmental determinants, such as gut microbiome composition, may also help explain the long-term sequelae of enteric infections. While there is evidence of differences in gut microbiome composition across different cultures, regions, and populations,[62–67] and environmental conditions,[68] to date specific WASH determinants of these differences, such as access to piped water, have not been evaluated using explicit counterfactuals. Thus measures of gut microbial conditions provide objective outcomes to more accurately measure the effect of WASH interventions[69,70] and capture long-term sequelae,[71] resulting in a more complete understanding of the health impacts.[72]

### 1.3. Overview of study

In the PAASIM study (*Pesquisa Sobre o Acesso à Água e a Saúde Infantil em Moçambique - Research on Access to Water and Children’s Health in Mozambique)*, we address a series of questions about the impact of community-level water system improvements on acute and chronic health outcomes in children in a low-income urban area of Mozambique. This matched-control cohort study follows mother-child dyads from late pregnancy through children 12 months of age, examining the impact of living in an area with an improved water network and/or having a household water connection on a variety of aspects of access to drinking water, microbes in the child gut (including both pathogens and other resident gut microbes), and ultimately downstream health outcomes (including diarrhea prevalence and growth) (**Figure 1**). This study will provide critical information for agencies who seek to understand how to optimize investments for improving child health, helping to fill the information gap about the impact of providing piped water to urban, low-income households by isolating the effects of major community-level water supply improvements on novel gastrointestinal disease outcomes.

**Figure 1.**
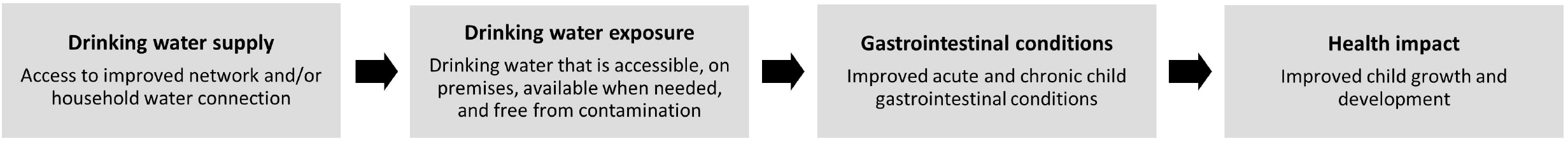
PAASIM theory of change. *No caption needed*

## 2. METHODS & ANALYSIS

### 2.1. Study setting and description of the intervention

Our study site is the coastal city of Beira, the second largest city in Mozambique (population ∼530,000),[73] which serves as a gateway for both the central interior portion of the country and a trade corridor to neighboring land-locked nations. The center of Beira is bordered by unplanned, informal settlements inhabited by over 300,000 low-income residents who often disproportionately feel the impact of severe weather due to lack of infrastructure.[74] A 2018 survey in Beira by Water & Sanitation for the Urban Poor (WSUP) showed that of 5,643 respondents, only 28% had a household water connection, and among those households without a connection, 83% used their neighbor’s tap as their main source of water (unpublished data, courtesy of WSUP). Therefore, improvement of water supply and delivery infrastructure is a priority.

The World Bank funded the Water Service & Institutional Support (WASIS-II) Project in 2016 to address the low access to improved water supply in Mozambique,[75] investing $140 million with the Mozambican public institutions FIPAG (responsible for the public and private investment program in urban water supply systems that serves as the water utility in Beira) and AURA, IP (the water regulatory authority responsible for the economic regulation and consumer protection of service provision). In addition, improvements in Beira are being augmented by investments from other groups, in particular the Dutch government, through infrastructure upgrades as well as emergency response funds following Cyclone Idai in 2019.[76] Improvements in the city of Beira include rehabilitation of water treatment facilities, replacing existing pipe mains that are failing, reticulation of water supply to new areas previously without water service, improving service in areas with poor coverage or low water pressure, and subsidizing water connection fees for the poor.

### 2.2 Study design

Several aspects of a city-wide water supply improvement project pose challenges to implementing a rigorous epidemiological study, particularly as people living in neighborhoods with piped water often differ in myriad ways from people living in neighborhoods without piped water. Water improvements to communities are often based on the needs or demographics of the community, government or donor priorities, and engineering considerations. Provisions of water supply to new areas previously without water service—or dramatic improvements in access and availability—represent a fundamental development that changes the livability and sometimes the makeup of the community. These issues lead to difficulty in finding a comparable control group for epidemiological comparison. Furthermore, rollouts of water interventions often happen in continuous phases over time, and these changes might coincide with other events or community improvements that similarly impact health, making it difficult for that community to serve as its own control in a pre post design.

By using a prospective matched cohort in this unique context of an ongoing natural experiment, we are able to overcome many of these difficulties. The prospective nature of our study allows better control of confounding through matching, restriction, and rigorous and thoughtful collection of potential control variables. We specifically focus on one region of the city (**Figure 2**), where some neighborhoods received water system improvements focused on preventing water losses through replacement of the distribution system pipe system in dense, low-income settings; other neighboring areas with similar demographic characteristics did not receive these improvements. Neighborhood-level matching took place in the context of a natural experiment, where the delayed rollout across the city allows us to find and compare intervention and control neighborhoods that are similar in many ways, before the rollout eventually reached all potential control areas.

**Figure 2.**
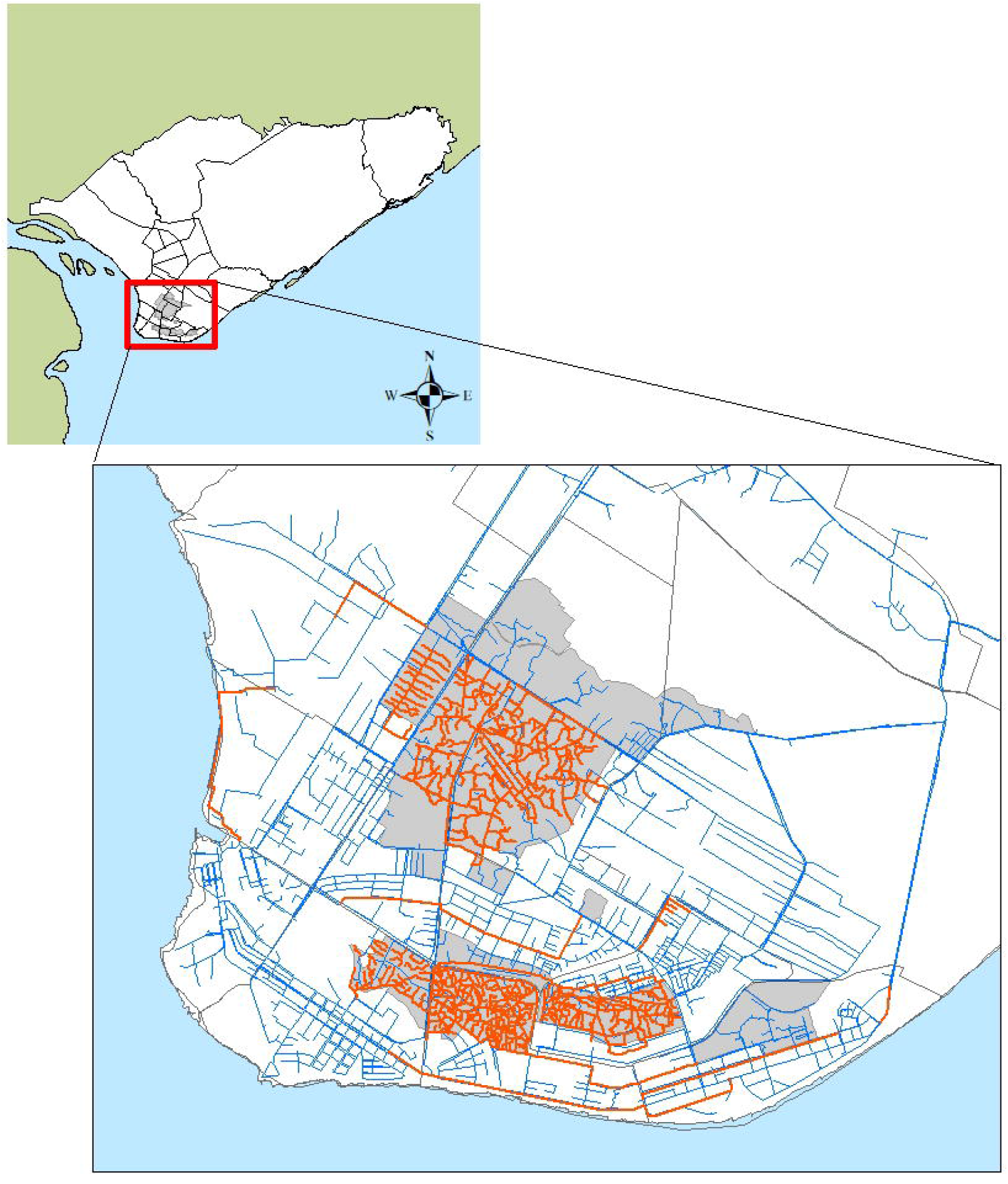
Map of PAASIM study site in Beira, Mozambique. Map of Beira, Mozambique, with enlargement highlighting study site. Red lines indicate the new distribution system water network. Blue lines indicate other parts of the water network. Gray shaded areas indicate neighborhoods enrolled in the study.

The Water Loss Reduction Project represents a subset of the improvements being carried out by FIPAG, with co-funding from the Dutch government and the World Bank WASIS-II project. The improvements are designed to reduce illegal connections, thereby increasing the water pressure and quality and increasing the system’s capacity for household connections. These areas also received some benefits related to improvements to the water intake and distribution systems. FIPAG undertook a campaign to offer new connections to households. These improvements were completed in some informal settlements in these low income areas of the city in 2019, with other adjacent neighborhoods with similar density and socioeconomic profile slated for completion in future years but not within the timeframe of our study. These specific distribution system upgrades therefore represent a unique opportunity to examine the impacts of community-scale water improvements with neighboring communities who did not receive the intervention serving as control areas for comparison. The neighborhoods under study are also in the lowest income—and therefore highest need areas of the city.

We will perform analyses that take into account the four factorial possible household types, based on mother-child dyad living in a sub-neighborhood with or without the improved water network and with or without a household connection (**Figure 3**). Our primary analyses focus on assessing A) the *total network effect*, by comparing subjects living in sub-neighborhoods with the improved water network (Household Types 1 and 2) to those living in sub-neighborhoods without these improvements (Household Types 3 and 4); and B) the *direct household connection effect*, by comparing subjects with household water connections on their premises (Household Types 1 and 3) to those without a connection (Household Types 2 and 4). Depending on the results of these two primary analyses, secondary analyses may evaluate the other comparisons depicted in **Figure 3**.

**Figure 3.**
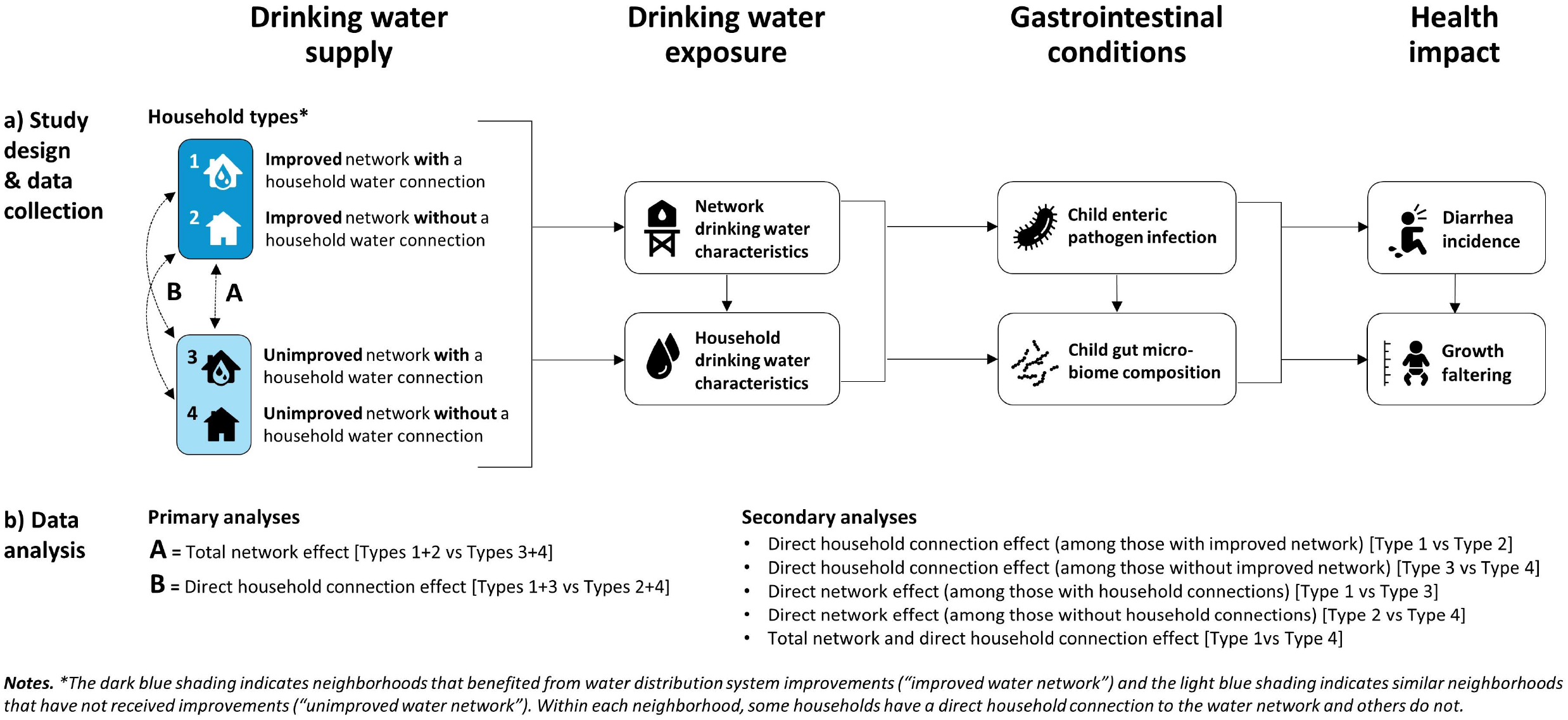
PAASIM study summary diagram. The diagram reflects the summarized a) study design and data collection, and b) data analysis approaches to isolate the effects of both overall water supply infrastructural improvements as well as the presence of a household water connection.

The reasons for some neighborhoods receiving the improvements and others not were a result of resource constraints. We conducted a population-based survey (described below) that allowed us to both restrict and match study sub-neighborhoods, thereby creating a statistically appropriate counterfactual for strong internal validity. The evaluation of a real-world intervention delivered in an informal urban setting provides strong external validity for estimating the effects of similar interventions in other low- and middle-income country urban sites. Our study design allows us to isolate the effects of both overall water supply infrastructural improvements as well as the presence of a household water connection. The presence of control areas not receiving upgrades adjacent to intervention areas that are matched on socioeconomic and density variables is unique to this study location. We collect data at multiple timepoints for each study household, allowing us to examine variability in each of the measures taken from each household, rather than at a single point in time, and also allowing for longitudinal analyses of the households and the individual enrolled subjects. We also employ rigorous measures of exposure and novel and objective outcome measures, including gut microbiome composition and molecular detection of enteric pathogens.

### 2.3. Patient & Public involvement

The executive secretary of AURA, IP was directly involved in the formation of the research questions, and FIPAG personnel were also engaged from the initiation of the project in helping develop the study design. Our team also received input from other public agency stakeholders during workshops that were held prior to initiation of the study. Study subjects and members of the general public were not involved in the study design. We provide regular updates with data summaries to public agency stakeholders, and plan to disseminate the main results to all study participants and also through public presentations for stakeholders in both Beira and Maputo.

### 2.4. Sub-neighborhood selection

Sub-neighborhood eligibility, selection, and matching of intervention and control sub-neighborhoods occurred through a two-step process:

#### 1) Intervention designation

Our study is a natural experiment, where the investigators had no control over the selection or timing of the intervention implementation. The study flow diagram is shown in **Figure 4**. We worked with FIPAG to determine which neighborhoods in Beira were to receive water distribution system upgrades prior to initiation of enrollment (2020) and before the end of the study (2022). FIPAG provided maps and timelines for construction works related to the upgrades, and the specific areas participating in the water loss reduction project. We also worked with FIPAG and through satellite imagery to identify similarly dense low income areas in Beira that were not slated to receive water network upgrades. A total of 17 potential neighborhoods were considered for inclusion in the study, and neighborhoods were divided into 80 sub-neighborhoods, delineated along natural boundaries such as roads or waterways. “Intervention” sub-neighborhoods include areas with the upgraded water distribution system. “Control” sub-neighborhoods include areas not receiving these improvements during the time period of the study. Within both intervention and control sub-neighborhoods, some households have a connection to the water system and others do not. We excluded nine sub-neighborhoods that were likely partially contaminated by proximity to the intervention or that were scheduled to receive the interventions within the timetable of our project; some control sub-neighborhoods are slated to receive the intervention after completion of our study.

**Figure 4.**
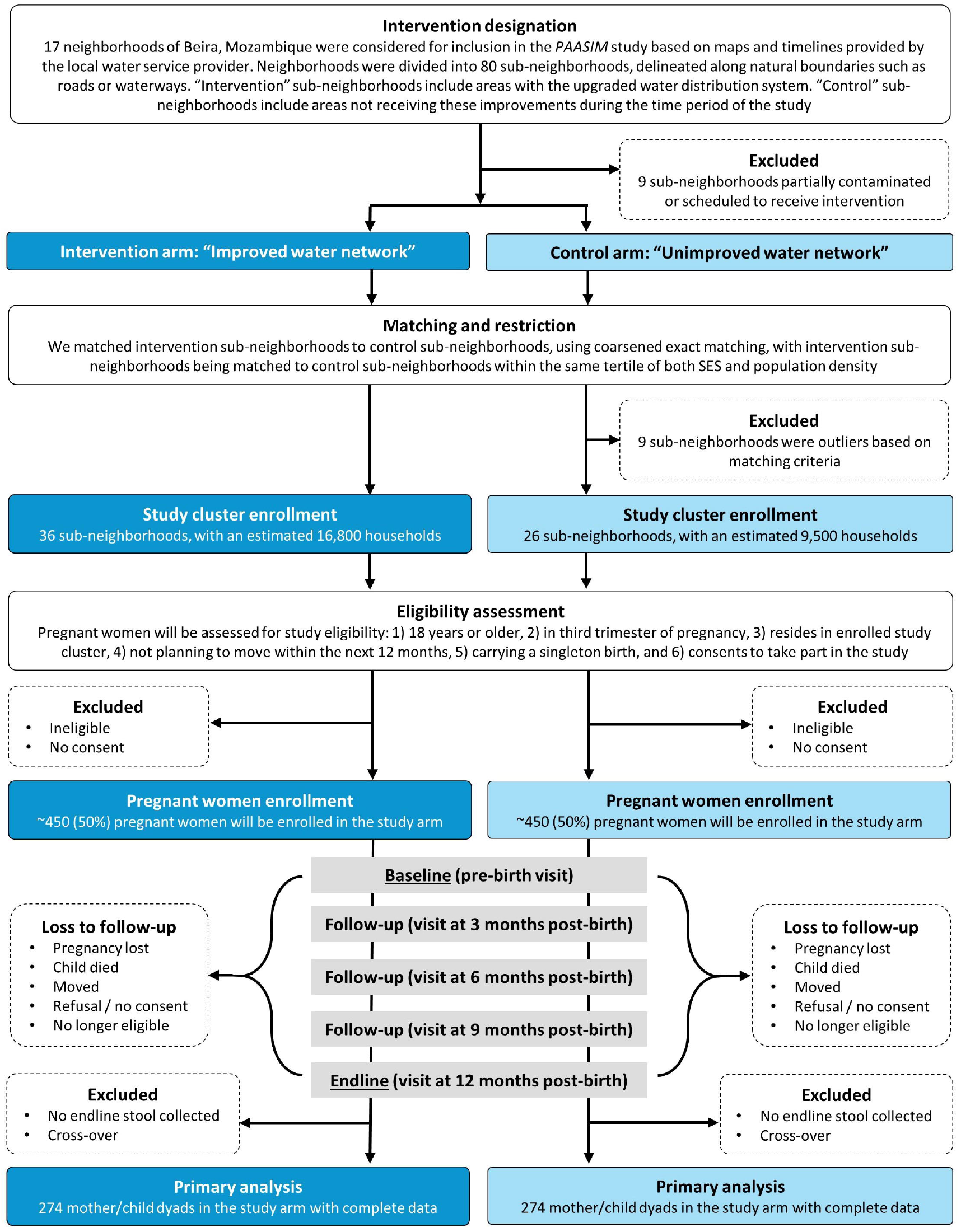
PAASIM study flow diagram. *No caption needed*

#### 2) Matching & Restriction

We followed the suggestion of Arnold et al. (2010) to use baseline (preintervention) data at the community level to match intervention to control communities when randomization is not possible.[22] To characterize subneighborhoods for further matching and restriction, we performed a population-based community survey in November-December, 2020 of approximately 1,700 households; this provided approximately a 5% proportional sample of our potential study sub-neighborhoods. We used a random grid sampling approach to estimate household density, using Google Earth satellite imagery, where a grid was placed over an area, and a random selection of squares were selected and counted independently in duplicate, and the number of houses per unit was extrapolated across unsampled squares. The survey contained modules regarding household demographics, water access and practices, sanitation access and practices, household assets and wealth indicators, as well as questions related to COVID-19. A socioeconomic status (SES) score was constructed using the ‘simple poverty scorecard’[77] developed specifically for Mozambique, and scores were aggregated at the sub-neighborhood level, and categorized into tertiles.

We matched intervention sub-neighborhoods to control sub-neighborhoods, using coarsened exact matching,[78,79] with intervention sub-neighborhoods being matched to control sub-neighborhoods within the same tertile of both SES and population density. Four neighborhoods (encompassing nine sub-neighborhoods) were found to be outliers in terms of their sub-neighborhood-level SES or sanitation, and were excluded from the study sampling frame. Ultimately we designated 36 intervention sub-neighborhoods, with an estimated 16,800 households, and 26 control sub-neighborhoods, with an estimated 9,500 households.

### 2.5. Participant recruitment, eligibility, and retention

We recruit pregnant women at the last trimester of pregnancy and follow the infant-mother dyads until the child is 12 months old (**Figure 5**). We selected the first 12 months of life because it is a critical development window,[80–82] it is a time when children are most at risk of acute and chronic effects of enteropathogen infection,[83] and it is a short enough period of time to avoid changes in water access that might occur. We recruit mothers at the end of their pregnancy so we can collect data on household risk factors (including drinking water quality) during the gestational period. Active recruitment occurs through identification of pregnant women in the 2020 population-based survey, lists of pregnant women visiting local health centers for pre-natal care, and study staff visiting under-enrolled sub-neighborhoods throughout the recruitment period. Based on Ministry of Health data for Sofala Province (where Beira is located), virtually all mothers attend pre-natal clinical visits.[84] Passive strategies include referrals of pregnant women by study participants and community leaders. We aim to have complete data on a total of 548 infant-mother dyads, approximately evenly divided between the intervention and control groups. We will continue to enroll dyads into both arms until we reach a minimum of 274 dyads with complete data in each arm, to ensure temporal balance throughout the duration of the study period.

**Figure 5.**
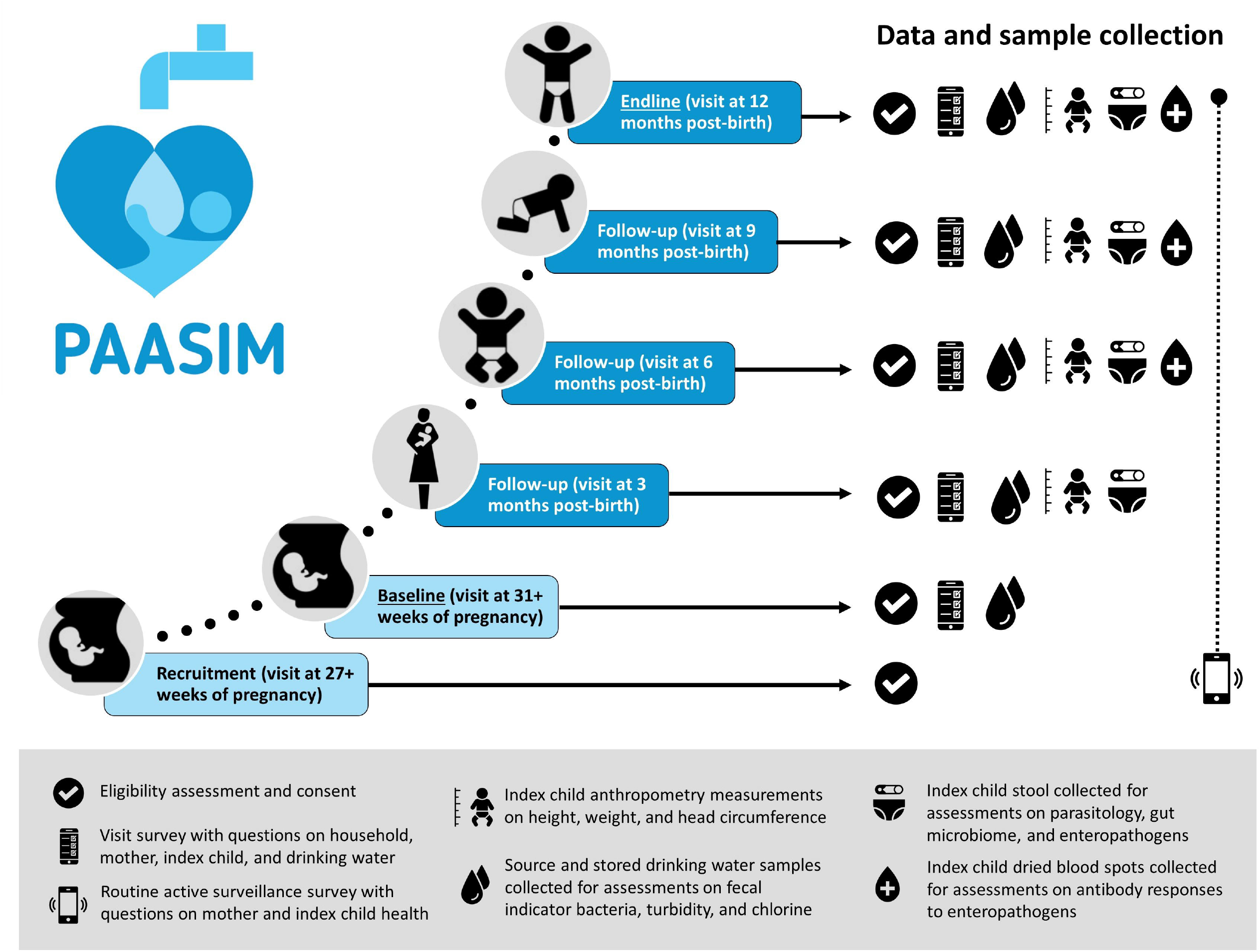
Data and sample collection timeline for outcomes in the PAASIM study. Data and sample collection of infant-mother dyads enrolled into the study will be used address a series of questions about the impact of community-level water system improvements on acute and chronic health outcomes in children in a low-income urban area of Mozambique.

During an initial pre-birth visit, pregnant women are assessed for study eligibility: 1) 18 years or older, 2) in third trimester of pregnancy, 3) resides in enrolled study cluster, 4) not planning to move within the next 12 months, 5) carrying a singleton birth, and 6) consents to take part in the study. We will re-assess study eligibility at each follow up visit and record if enrolled participants have been lost to follow up.

### 2.6. Data collection

A local data collection firm (WE Consult) performs the in-country coordination of participant enrollment, data collection, and sample collection. Enumerators conduct household visits before birth for consent, eligibility, and conditions. At months 3, 6, 9, and 12 we deploy survey instruments to collect data on key indicators through structured observations, reports from respondents, and objective measurements (**Table S1**). We assess a number of variables related to drinking water, including aspects of water quality, water access, water availability, water security, water consumption, and participant satisfaction with water. Brief active surveillance calls also take place monthly by phone to gather information on prenatal and perinatal environmental exposures and illnesses (**Figure 5**). We ask the caregiver to report diarrhea and blood in the stool (dysentery) of the index child in the previous week at the 3, 6, 9, and 12 month surveys and during active surveillance calls; due to concerns about reporting biases, we also include negative control outcomes.[85] At each post-birth visit we measure child: 1) length, weight, and head circumference, and calculate length-for-age and weight-for-age Z-scores. Prevalence of stunting and underweight are defined as two standard deviations below median of the reference population.[86] All data are collected on electronic tablets using Open Data Kit (ODK) Collect, an open-source program which allows offline data collection on a mobile device.[87] Additional details are provided in the Supplementary Material.

### 2.7. Sample collection, processing, and analysis

We briefly describe sample collection and downstream processing and analysis here, with additional details provided in the Supplementary Material.

#### 2.7.1. Stool

Stool of the index child is collected at months 3, 6, 9, and 12. Three aliquots are placed in temperature stable lysis buffer collection tubes, and two additional aliquots are used to prepare a slide for Kato-Katz analysis of parasite ova[88]. Eligible participants are referred for deworming medicine at the 12-month visit, after returning results of the parasitological exam to study subjects in collaboration with Instituto Nacional de Saúde (INS) staff in Beira.

Extracted nucleic acids are analyzed: (1) using the TaqMan Array card (TAC, ThermoFisher Scientific, Waltham, MA, USA) assay, which allows quantification by real-time PCR via a 384-well microfluidic card for simultaneous detection of multiple viral, bacterial, and parasitic enteric pathogen targets as well as antimicrobial resistance genes,[89] customized for our targets of interest (**Table S2**); and (2) by sequencing of the V4 region of the 16S ribosomal RNA (rRNA) gene amplicon to characterize gut microbiome community structure and composition. Bioinformatic analyses will be completed using the QIIME2 software platform.[90]

#### 2.7.2. Dried blood spots

##### Sample collection

A trained nurse or laboratory technician collects up to six dried spots of capillary blood of the index child at 6, 9, and 12 month visits on Tropbio Filter Paper Blood Collection Disks (Cellabs, Sydney, Australia), using a 2mm lancet. Samples are stored at -20 °C and shipped at ambient temperature.[91] We will use the Luminex platform to carry out high throughput, multiplex antibody assays that enable the simultaneous measurement of quantitative antibody responses to dozens of pathogens from a single blood spot.[92] Our first measure will occur at 6 months, to avoid detection of maternal antibodies that wane over the first 3-6 months of life.[93]

#### 2.7.3. Drinking water

##### Sample collection

We collect 100-mL household drinking water samples from source and stored water at all household visits. To complement the household sampling, we collect samples from a selection of 45 public standpipes located within the study area and 55 additional public standpipes located elsewhere in the city of Beira. At public standpipes we also measure water pressure by measuring time to fill a fixed volume (1L or 5L, depending on the pressure). Samples are processed for fecal indicator bacteria within six hours of collection using Colilert-18 reagent and the Quanti-Tray/2000 MPN method (IDEXX Laboratories, Westbrook, ME, USA), as well as for free and total chlorine levels and additional physiochemical parameters (pH, conductivity, and turbidity). Large volume samples will be collected from a subset of 50 households (1 L, processed by membrane filtration) and 25 public standpipes (50 L, processed by dead end ultrafiltration[94]) in two different seasons, and tested for enteropathogens using the TAC assay.

### 2.8. Outcomes

Our primary outcomes include: any bacteria or protozoa infection at age 12 months after birth; individual pathogens or pathogen groups; child gut microbiome composition; and household source water quality. While we measure viral pathogens using the TAC assay, they will be excluded from the combined enteropathogen prevalence primary outcome measure, because waterborne transmission is unlikely to dominate for these viral pathogens.[95–98] In addition to the aforementioned reasons related to child development and infection risk, measuring pathogens at 12 months will give us the greatest power to detect a difference, given higher levels of infection at that age than in younger children. We will measure gut microbiome using 16S rRNA gene amplicon sequencing in the full sample at 12 months and in a random subset of 200 children with complete data at 3, 6, and 9 months, evenly distributed between intervention and control groups; dyads eligible for sub-set sampling will include those with complete stool sample collection and unchanged intervention exposure conditions. The 12-month samples will allow us to compare all study children at a common time, when all children are consuming drinking water and once the gut microbiome has become relatively established;[99] the longitudinal samples will allow for comparison of development of the microbiome over time between the two groups. Microbiome outcomes include alpha and beta diversity metrics, and identification of enriched taxonomic groups. We also include household source water quality as a primary exposure outcome, as understanding whether exposure to microbial contaminants is altered is considered a critical aspect of evaluation of WASH projects.[70,100]

Additional non-primary outcomes include pathogen count, pathogen community similarity (measured using Jaccard similarity index), diarrhea, child growth, and prior enteropathogen infection (measured using serology on dried blood spot samples). We will measure additional water quality exposure measures, as well as measures of exposure to the improved water system, such as fidelity of the intervention (e.g., improvements to water quantity and coverage of household taps) and receipt of the intervention by community members (e.g., reductions in water insecurity, increased water use). These fidelity and uptake measures will be collected at all time points through direct observation and respondent report. Available minimal detectable effect sizes are summarized in **Table 1** and calculations are further detailed in the Supplementary Material.

**Table 1.** Primary and non-primary health outcomes and exposure outcomes for the PAASIM Study. Patent enteric pathogen infection in stool is measured via TaqMan Array Card (TAC) assay; Stool microbiome composition is measured via 16s rRNA amplicon sequencing; prior enteric pathogen exposure is measured via serological assays of dried blood spots. Drinking water quality measured by IDEXX as *E. coli* most probable number/100mL. See text for further details. Calculations and additional values for the minimum detectable effect are described in the Supplementary Material.

|  | Time point | Minimum detectable effect (Risk Ratio) | Anticipated control group prevalence<sup>a</sup> |
| --- | --- | --- | --- |
| **Primary outcomes** |  |  |  |
| Patent enteric pathogen infection |  |  |  |
| Prevalence of any bacterial or protozoan pathogen | 12 mo<sup>b</sup> | 0.74 | 70% |
| Prevalence of any bacterial pathogen | 12 mo<sup>b</sup> | 0.69 | 61% |
| Prevalence of any protozoan pathogen | 12 mo<sup>b</sup> | 0.50 | 32% |
| Any co-infection (bacterial, protozoan, or viral pathogens) | 12 mo<sup>b</sup> | 0.60 | 45% |
| Gut microbiome composition | 12 mo<sup>c</sup> |  |  |
| Alpha diversity |  | -- | NA |
| Beta diversity |  | -- | NA |
| Enriched taxa |  | -- | NA |
| Household source drinking water quality | 12 mo<sup>d</sup> | -- | NA |
| **Additional health outcomes** |  |  |  |
| Patent enteric pathogen infection |  |  |  |
| Pathogen count | 12 mo<sup>b</sup> | -- | -- |
| Pathogen community similarity | 12 mo<sup>b</sup> | -- | -- |
| Individual pathogens | 12 mo<sup>b</sup> | n/a-0.49 | 2-31% |
| Any virus | 12 mo<sup>b</sup> | 0.46 | 28% |
| Gut microbiome composition | 3, 6, 9 mo<sup>c</sup> |  |  |
| *(Same variables as gut microbiome composition for 12 mo)* |  |  |  |
| Prior enteric pathogen exposure | 6, 9, 12 mo | -- | -- |
| *(Same variables as patent enteric pathogen infection)* |  |  |  |
| Diarrhea 1-week period prevalence (caregiver report) | Weekly | 0.26 | 14.4% |
| Anthropometric Measurements | 3, 6, 9, 12 mo |  |  |
| Length-for-age Z-score |  | -- | -- |
| Weight-for-age Z-score |  | -- | -- |
| Stunting prevalence<sup>c</sup> |  | 0.49 | 31% |
| Underweight prevalence<sup>c</sup> |  | 0.22 | 13% |
| All-cause mortality (while enrolled in study) | Continuous | -- | -- |
| **Additional Exposure outcomes** |  |  |  |
| Primary drinking water source | 3, 6, 9, 12 mo |  |  |
| Drinking water quality (source) | 3, 6, 9 mo<sup>f</sup> |  |  |
| Drinking water quality (stored) | 3, 6, 9, 12 mo |  |  |
| Water access | 3, 6, 9, 12 mo |  |  |
| Water availability | 3, 6, 9, 12 mo |  |  |
| Water security | 3, 6, 9, 12 mo |  |  |
| Water consumption | 3, 6, 9, 12 mo |  |  |
| User satisfaction with water | 3, 6, 9, 12 mo |  |  |

<sup>a</sup>Anticipated control group prevalence based on control group prevalence for 10-14 month olds in the MapSan trial ([31] and J. Knee, pers. Comm.).
<sup>b</sup>Samples also collected at 3, 6, and 9 months of age may also be analyzed, depending on results of primary analysis at 12 months.
<sup>c</sup>A subset of n=200 samples will be analyzed for gut microbiome composition in children at 3, 6, and 9 months of age. All 12 month samples will be analyzed.
<sup>d</sup>This is a conservative estimate as it does not account for weekly active surveillance.
<sup>e</sup>Defined as two standard deviations below median of the reference population.
<sup>f</sup>Samples will be analyzed at 3, 6, 9, and 12 mo, but 12 month samples are the primary outcome of interest.

### 2.9. Analysis plan

The pre-analysis plan for this study is published on the Open Science Framework platform (https://osf.io/4rkn6/).

#### 2.9.1. Total network effect

To assess the impact of the intervention on our primary enteric pathogen infection outcomes and water quality exposure outcome (**Table 1**), we will use an intention-to-treat (ITT) analysis approach to compare children living in intervention versus control sub-neighborhoods, without regard to uptake/use of the intervention (i.e., direct household connection on the premises). We will use multivariable log-linear binomial regression models, as pathogen infection is a binary variable, and will use generalized estimating equations (GEE) to account for clustering at the sub-neighborhood level. We group matched on sub-neighborhood-level SES and population density, using weighting to account for unequal numbers between the intervention and control areas within each matching stratum.[101] We will additionally control for household- and individual-level confounders, including household SES, household sanitation, mother’s education-level, and child sex. We may adjust for additional variables if there are found to be imbalances in potential confounders in our baseline assessment. We hypothesize that the intervention will lead to reductions in enteric pathogens among children and microbial water contamination of source water.

For additional outcomes and exposure variables of interest (**Table 1**), we will use a similar modeling approach, using log-linear binomial regression models for binary outcomes, linear regression models for continuous outcomes, and Poisson (or negative binomial) models for count outcomes. For outcomes measured at multiple time points, we will present results separately for each given time point. For these analyses, we will control for sub-neighborhood-level SES and population density through matching, and will additionally control for household sanitation, mother’s education-level, child sex, and any other variables that are imbalanced and are conceivably potential confounders. For previous enteropathogen exposure evaluated using serological measures we hypothesize that those in the intervention group will show delays in pathogen acquisition.

To assess the impact of the intervention on microbiome outcomes, we will evaluate alpha diversity (Chao1 species richness estimator, Pielou’s *J* evenness estimator, and the Shannon diversity index[102]) using the same modeling approach as described above for continuous outcomes. Linear discriminate effect size (LEfSe) analyses will be used to evaluate specific 16S rRNA gene-based Operational Taxonomic Units (OTUs) that differ between individuals in intervention versus control groups, and will include effect size corrections[103]. We will examine the impact of intervention group, controlling for other covariates, on community similarity using Adonis permutation models,[104] based on weighted UniFrac and Bray-Curtis distances, and evaluate and visualize differences using PCA and/or NMDS plots. We hypothesize that we will be able to observe detectable differences in gut microbiome composition in children living in intervention versus control sub-neighborhoods and we will report these differences at the individual OTU and bacterial family levels.

#### 2.9.2. Direct Household Connection Effect

To assess the effect of having a water connection at the household or compound, we will use models similar to those described above, but accounting for a household network connection. We will also assess the interaction between the household and neighborhood network variables, which will allow us to contrast and estimate indirect, direct, and total effects, as shown in **Figure 3**. We hypothesize that participants with both improved water networks in their sub-neighborhoods and household water connections will most benefit from the interventions in terms of our primary and non-primary health outcomes and exposure outcomes of interest.

#### 2.9.3. Additional analyses

For select primary outcomes, we will assess if there is effect modification by a third variable, such as follow-up round/age, participant sex, and household sanitation access. We will use interaction terms to identify potential interactions, and will present stratified results (e.g., separately by sex) if interactions are detected. The intervention status of subneighborhoods was set at baseline, but if control subneighborhood(s) receive the intervention after the study has started, we will perform sensitivity analyses dropping and/or recategorizing subneighborhood(s) that crossed over.

There are several analyses where we do not use the matched-design and intervention variable in our analyses. For example, we will assess associations between various water measures on health, without regard to the intervention designation. We will also examine changes in the gut microbiomes of children over time. Additional analyses will be described and documented in OSF.

### 2.10. Sample size and power calculations

Our minimal sample size of 548 households–half in intervention and half in matched control sub-neighborhoods–was powered for our primary outcome of prevalence of any non-viral pathogen. Utilizing data from the MapSan trial for children 10-14 months of age (J. Knee, *pers comm*) we used a control group prevalence of 70% for any non-viral pathogen, and estimated the ability to detect a relative risk of 0.74, alpha=0.05, and power=80% using a two-sided test for significance.[31] We estimated a sub-neighborhood-level interclass correlation (ICC) of 0.05 (a moderate estimate) among our 62 designated sub-neighborhoods. We will also report on the final ICC and other assumptions of this power analysis at the end of the study. Estimates of minimum detectable effect sizes based on control prevalence of the outcome of interest (**Table S2**) show we may be adequately powered to detect a difference in some individual pathogens if those pathogens have high prevalence and/or if they are strongly associated with the water supply improvement intervention (e.g., waterborne pathogens). We target planned recruitment at 900 pregnant women in the third trimester, to account for incomplete data and loss to follow-up. We used sub-neighborhood enrollment targets proportionate to our density estimates to achieve balance across intervention and control sub-neighborhoods.

### 2.11. Blinding

All lab personnel and field enumerators are blinded to the intervention status of the samples and households. Participants cannot be blinded to their household-level water exposure status or cluster-level exposure status, although participants may or may not know about water improvements in their particular neighborhood. A primary analyst external to the core data management team is blinded to the group assignments until the data cleaning and primary analysis are completed. Details of these procedures are included in the Supplementary Material. Unblinding will occur only after primary outcome models are developed and compared between two independent analysts. Analyses examining the impact of the intervention on non-primary outcomes or exposures of interest will not be unblinded until after analyses that examine the impact of the intervention on our primary outcomes have been completed. Purely observational analyses that do not require information on intervention group may be completed before unblinding occurs.

## 3. ETHICS & DISSEMINATION

The study protocol, informed consent forms, and data collection tools were approved by 1) Mozambique National Bio-Ethics Committee for Health (IRB00002657) and (2) Emory University’s Institutional Review Board (IRB00098584). As this study is a natural experiment that the investigators do not control, we do not have a data monitoring committee or any interim stopping guidelines. Enrollment for this study began during the COVID-19 pandemic, and precautions were taken to secure the safety of study staff and participants based on guidance from INS, Emory University, and the University of Washington.

Any changes to this published protocol will be noted in OSF, and, where relevant, in future publications. De-identified data sufficient to replicate study findings will be publicly available on OSF upon completion and publication of the study results. A report will also be prepared and shared with the municipality and health authorities in Beira, and other relevant stakeholders. All microbial DNA sequence data will be made available through the SRA database of NCBI upon validation and/or publication of the corresponding manuscript.

## Data Availability

De-identified data sufficient to replicate study findings will be publicly available on OSF upon completion and publication of the study results. All microbial DNA sequence data will be made available through the SRA database of NCBI upon validation and/or publication of the corresponding manuscript.

https://osf.io/4rkn6/

## AUTHORS’ CONTRIBUTIONS

MM and MCF conceived of the overarching idea of evaluating the intervention; KL and MCF conceived of the specific study and secured funding; JVG and MCF designed the analysis plan, with input from LW; ZAC, SH, SM, JSS, and MKMP designed protocols for recruitment of participants and oversaw collection of field data, with guidance from RN and JLM; JSS and JVG oversaw data management, with help from SH, MKMP, SM, and CSFS; CV and CSFS designed specimen management and laboratory protocols; TC, JB, and LW advised on study design, epidemiological approaches and research methods; RN, MM, and JLM provided oversight on relevant scientific questions in Mozambique; RN, JSS, CV, KL, MCF, ZAC, SM, and MKMP managed human subjects protocol submissions; KK and KL oversaw microbiome analysis approach; RN oversaw parasitology analysis approach; KL and JB oversaw enteric pathogen analysis approach; KL and JVG wrote the manuscript, with input from all authors.

