## Supplemental materials for "Study Design and Rationale for the PAASIM Project, a Matched Cohort Study on Urban Water Supply Improvements and Infant Enteric Pathogen Infection, Gut Microbiome Development, and Health in Mozambique"

**Supplementary Material**

### Details of data collection

A data collection firm (WeConsult) performs the in-country coordination of participant enrollment, data collection, and sample collection. Enumerators are recruited from Beira; all are fluent in Portuguese (spoken by >80% of the study population)[1] and more than half speak the local language. Enumerators conduct household visits for consent and eligibility pre-birth, and at months 3, 6, 9, and 12 post-birth; active surveillance calls take place monthly (**Figure 4**). All data are collected on electronic tablets using Open Data Kit (ODK) Collect, an open source program which allows offline data collection on a mobile device.[2]

#### *2.4.1. Household visit*

Survey instruments consist of several modules aimed at collecting data on key indicators through structured observations, reports from respondents, and objective measurements (Table S1). The survey includes household questions (e.g., SES, demographics, nutrition and food insecurity, flooding, animals, and sanitation and handwashing facilities), questions about the mothers (e.g., demographics, medical and physical health, pregnancy and birth care, and breastfeeding practices), questions about the index child (e.g., birth outcome, nutrition, health and recent illness, anthropometry, and vaccination history), and household questions about drinking water. We also collect GPS coordinates at each household visit.

#### *2.4.2. Active surveillance calls*

A brief monthly active surveillance call is conducted by phone to gather information on prenatal and perinatal environmental exposures and illnesses, on child illness symptoms, and intake of medicines, vitamins, breastfeeding, and introduction of complementary foods. The first active surveillance pre-birth call is targeted to occur one month after enrollment and calls continue on a monthly basis between visits. To facilitate communication with the study team participants receive a 150 MZN phone credit at each visit.

#### *2.4.3. Drinking water*

Drinking water characteristics, which are on the causal pathway between the intervention and our study outcomes (**Figure 1**), will be used to provide evidence of biological plausibility of the intervention's effects on these downstream outcomes. We will characterize water access using the Joint Monitoring Program definition of "safely managed," i.e., an improved water source that is accessible on premises (located within the dwelling, yard or plot), available when needed (sufficient water available for at least 12 hours per day), and free from contamination (no *E. coli* detected in a 100mL sample).[3,4] We also assess a number of additional variables related to drinking water, including aspects of water quality, water access, water availability, water security, water consumption, and participant satisfaction with water. These variables are evaluated throughout the study at the household level and also at the community level through objective, observed, and participant-reported measures.

#### *2.4.4. Diarrhea*

At months 3, 6, 9, and 12, we ask the caregiver to report diarrhea and blood in the stool (dysentery) of the index child in the previous week. We use the case definition of diarrhea as 3 or more loose stools in a 24 hour period.[5] Due to concerns about reporting biases, we also include a negative control outcome[6]; caregivers report on accidents that resulted in physical injury in the previous week. In addition, we note objective characteristics of the stool samples collected, including observed blood and mucus and an infant stool form scale describing consistency (4-point scale), amount (4-point scale), and color (6 categories).[7]

#### *2.4.5. Anthropometry*

At months 3, 6, 9, and 12, we measure child: 1) length, weight, and head circumference. From these measures we will calculate Z-scores (LAZ and WAZ), and prevalence of stunting and underweight, defined as two standard deviations below median of the reference population.[8] We measure weight using digital baby weighing scales with 5g gradations (ADE, Model# M112600, Hamburg, Germany), and we measure height using a baby length measuring board with 1mm gradations (ADE, Model# MZ10040).[9] Measurements are repeated twice and recorded into the survey. The enumerator is prompted to conduct a third measurement if there are differences of >1 cm in length or head circumference or if weight is off by >0.5kg in the two repeated measurements. Each child's LAZ and WAZ will be calculated using the WHO Child Growth Standards for the reference population[10,11] and WHO Anthropometric macros.[12]

**Details of sample collection, processing, and analysis**

*2.5.1. Stool*

*Sample collection:* Stool of the index child is collected at months 3, 6, 9, and 12 using a diaper, which is provided to the primary caregiver before a household visit. If a fecal sample is not provided during the initial home visit, we leave the primary caregiver with additional diapers and two more attempts are made to collect the sample within 4 hours of production. If needed, we provide a cooler and cold-pack, and collect the sample within 7 hours of production.

*Sample processing:* Lab personnel transfer 1g fecal material from the diaper into a DNA/RNA Shield Fecal Collection Tube (Zymo Research, Irvine, CA, USA); we aim to collect three aliquots in separate collection tubes. The DNA/RNA Shield stabilization buffer lyses the cells and renders the sample DNA and RNA stable for 30 days at ambient temperature.[13] Samples are stored and shipped at -20 °C. Nucleic acids are extracted from fecal samples using the QIAamp 96 Virus QIAcube HT Kit on a QIAcube HT (Qiagen Sciences Inc., Germantown, MD, USA), and stored at -80°C until further processing.

*Analysis - parasites*: Immediately after the first three aliquots are placed in the collection tubes, two additional aliquots of fecal material are taken from the diaper to prepare a slide for Kato-Katz analysis of parasite ova[14]. If sufficient material is available for a second slide analysis is carried out in duplicate. Samples are analyzed for hookworm (*e.g., Necator americanus*, *Ancylostoma duodenale*) immediately following slide preparation, and for *Ascaris* spp., *Schistosoma mansoni*, *Trichuris trichiura*, *Taenia*, *Enterobius vermicularis*, and *Strongyloides stercoralis* after overnight incubation at room temperature. Eligible participants are referred for deworming medicine at the 12-month visit, after returning results of the parasitological exam to study subjects in collaboration with Instituto Nacional de Saude (INS) staff in Beira.

*Analysis - enteric pathogens:* Extracted nucleic acids are analyzed using the TaqMan Array card (TAC, ThermoFisher Scientific, Waltham, MA, USA) assay, which allows quantification by real-time PCR via a 384-well microfluidic card for simultaneous detection of multiple viral, bacterial, and parasitic enteric pathogen targets as well as antimicrobial resistance genes. Immediately prior to nucleic acid extraction, samples used in downstream TAC assays are seeded with the Inforce 3 Bovine Vaccine (Zoetis, Parsippany-Troy Hills, NJ, USA)[15] containing Bovine Herpesvirus 1 (BHV) and Bovine Respiratory Syncytial Virus (BRSV) as extrinsic controls, to monitor extraction and amplification efficiency. Pathogens will be linked to the diarrheal disease episode based on relative cycle threshold values from the TaqMan results.[16] The specific targets we will test for are shown in **Table S2**. The batch of TaqMan Array Cards will be QA/QC using positive control plasmids. Initially, a standard curve for quantification will be run in duplicate, and the limit of detection and limit of quantification will be determined by running cards at a low concentration (concentration determined based on the standard curve tests) until a 95% positivity rate out of at least ten assays is obtained. Additionally, when processing samples, standard curves will be run in singlicate once per month with the positive control plasmid. A negative control will be run per card.

*Analysis - Gut microbiome composition:* We will characterize the gut microbial community structure and composition by sequencing of the V4 region of the 16S ribosomal RNA (rRNA) gene amplicon. Immediately prior to nucleic acid extraction, samples used in downstream 16s assays will be seeded with the ZymoBIOMICS Spike-in Control I (High Microbial Load) (Zymo Research) containing *Imtechella* *halotolerans* and *Allobacillus halotolerans*. Bioinformatic analyses will be completed using the QIIME2 software platform and other bioinformatics tools[17].

*2.5.2. Dried blood spots*

*Sample collection:* A trained nurse collects up to six dried spots of capillary blood of the index child at 6, 9, and 12 month visits on Tropbio Filter Paper Blood Collection Disks (Cellabs, Sydney, Australia), using a 2mm lancet.

*Sample processing:* Samples are allowed to dry overnight, then three aliquots of two spots each are placed in a Ziploc bag with silica desiccant. Dried blood spots can be stored at ambient temperatures for up to 100 days, even in tropical climates[18,19] but samples are stored at -20 °C and shipped at ambient temperature.[20]

*Analysis – antibodies*: We will use the Luminex platform to carry out high throughput, multiplex antibody assays that enable the simultaneous measurement of quantitative antibody responses to dozens of pathogens from a single blood spot.[21] Bead coupling of antigens will occur at the U.S. Centers for Disease Control and Prevention (CDC), and CDC collaborators will also provide support in determining appropriate antigen cut points. Our panel includes a subset of enteropathogens that have targets on the TAC assay, including *Giardia*, *Cryptosporidium*, *Entamoeba histolytica*, norovirus, *Campylobacter*, enterotoxigenic *E. coli* and *V. cholerae,* following previous similar studies.[22] Our first measure will occur at 6 months, to avoid detection of maternal antibodies that wane over the first 3-6 months of life. [23]

*2.5.3. Drinking water*

*Sample collection:* We collect 100-mL household drinking water samples from source and stored water at all household visits. We select the drinking water source sample by asking the mother what the source of water is for the household that would be given to the study child to drink, or to mix formula. If the source water is not in the household, the enumerator walks with the mother to the source (e.g., a neighbor's house or public standpipe) to collect a sample and GPS location. We select the stored water sample by asking the mother for any water in the household that is used for drinking purposes; stored water is water stored in a jerry can, bucket, or other container in the household for later consumption.

To complement the household sampling, we also collect samples from a selection of 45 public standpipes located within the study area and 55 additional public standpipes located elsewhere in the city of Beira. At public standpipes we also measure water pressure by measuring time to fill a fixed volume (1L or 5L, depending on the pressure).

*Sample processing and analysis:* All samples are placed on cold packs after collection for transport to a lab in Beira. Samples are processed for fecal indicator bacteria within six hours of collection using Colilert-18 reagent and the Quanti-Tray/2000 MPN method (IDEXX Laboratories, Westbrook, ME, USA). Free and total chlorine levels are measured using a DR300 Pocket Colorimeter (Hach Company, Loveland, CO, USA) and DPD powder pillows (Hach Company)[24]. Additional physiochemical parameters (pH, conductivity, and turbidity) are measured for public standpipe water samples using a Pocket Pro+ Multi 2 Tester and TL2300 turbidity meter (Hach Company).

**Details of Blinding**

At enrollment, households are assigned a unique identifier independent of intervention status. The core data management team conducts quality assurance using geocoded data to ensure group exposure status aligns with cluster-level designations of the exposure (i.e., intervention vs control areas). All geocoded data and group exposure indicators are removed prior to an external analyst performing the data cleaning and primary analysis. Data are cleaned, including decisions on missing data, outliers, and variable categorizations, before the analyst receives any group exposure. The analyst performs the primary analyses making comparisons between undefined group exposures. Once the analysis models are finalized, these rehearsal results are input into tables to create table shells for the final analyses. The primary analyst then receives a masking key, with true cluster-level designations of the exposure, and reruns the code with the appropriate group exposure, and re-inputs the final results into the table shells.

**Table S1. Household survey data collection in the *PAASIM* Study**

| Category | Indicator | Time point | | | | | Type of data |
|---|---|---|---|---|---|---|---|
| | | Pre-birth | 3mo | 6mo | 9mo | 12mo | |
| **Household-level** | | | | | | | |
| **General characteristics** | Socio-economic status | X | * | * | * | X | Respondent reported<br>Observation of households |
| | Demographics (household ownership, household members, children under 5 years of age, primary wage earner) | X | * | * | * | X | Respondent reported |
| **Nutrition** | Food insecurity | X | X | X | X | X | Respondent reported |
| | Types of food consumed by household members yesterday | | | | | | Respondent reported |
| **Sanitation** | Access to improved sanitation | X | X | X | X | X | Observation of household latrines |
| | Location of sanitation facility | X | X | X | X | X | Observation of household compounds |
| | Sharing of sanitation facility | X | X | X | X | X | Respondent reported |
| | Sanitation facility characteristics (serviceable, drop hole cover, smooth and cleanable floor) | X | X | X | X | X | Observation of household latrines |
| | Presence of human feces in the household compound | X | X | X | X | X | Observation of household compounds |
| | Trash disposal | X | X | X | X | X | Respondent reported |
| **Handwashing** | Access to handwashing facility with soap and water | X | X | X | X | X | Observation of household compounds |
| **Flooding** | Flooding of household compound | X | X | X | X | X | Respondent reported |
| **Animals** | Presence of animals in the household or compound (chickens, ducks/turkey, dogs, cats, pigs, sheep, goats, rabbits, donkeys) | X | X | X | X | X | Respondent reported |
| | Presence of animal feces in the household compound | X | X | X | X | X | Observation of household compounds |
| | Child(ren) contact with animals | X | X | X | X | X | Respondent reported |
| **Health** | Deworming history (pre-school age children, school age children, and mother) | | X | X | X | X | Respondent reported |
| **Moving** | Moved to another household | | X | X | X | X | Respondent reported<br>Observation of household compounds |

Notes: *Asked if study household has moved to another location within the study area

| Category | Indicator | Time point | | | | | Type of data |
| --- | --- | --- | --- | --- | --- | --- | --- |
| | | Pre-birth | 3mo | 6mo | 9mo | 12mo | |
| **Mother-level** | | | | | | | |
| **General characteristics** | Demographics (age, religion, language, education, employment, marital status) | X | | | | X | Respondent reported |
| **Mental and physical health** | WHO 5 well-being index | X | X | X | X | X | Respondent reported |
| | Medical diagnosis history (diabetes, high blood pressure, heart disease, urinary tract infection, kidney disease, sexually-transmitted disease, cancer, malaria) | X | | | | | Respondent reported |
| | Vaginal health practices | X | | | | | Respondent reported |
| **Pregnancy health** | Pregnancy history | X | | | | | Respondent reported |
| | Expected delivery date | X | | | | | Respondent reported |
| | Pre-natal care | X | | | | | Respondent reported |
| | Medical diagnosis during pregnancy (gestational diabetes, high blood pressure, placenta previa, COVID-19, Dengue, Zika, Chikungunya, Malaria) | X | | | | | Respondent reported |
| | Medications, vitamins, or supplements during pregnancy | X | | | | | Respondent reported |
| **Birth care** | Delivery location | | X | | | | Respondent reported |
| | Cesarean section | | X | | | | Respondent reported |
| | Post-natal care | | X | | | | Respondent reported |
| **Breastfeeding** | Breastfeeding intentions | X | | | | | Respondent reported |
| | Breastfeeding practices | | X | X | X | X | Respondent reported |
| **Sanitation** | Primary place of defecation in last week | X | X | X | X | X | Respondent reported |
| | Exclusive use of sanitation facility in last week | X | X | X | X | X | Respondent reported |
| **Travel** | Estimated time spent away from home in the last 12 months | | | | | X | Respondent reported |

| Category | Indicator | Time point | | | | | Type of data |
|---|---|---|---|---|---|---|---|
| | | Pre-birth | 3mo | 6mo | 9mo | 12mo | |
| **Child-level** | | | | | | | |
| **Birth** | Birth outcome | | X | | | | Respondent reported |
| | Birthdate | | X | | | | Observation of child health card |
| | Birth weight | | X | | | | Respondent reported |
| | Sex | | X | | | | Observation of child health card |
| **Health** | Mortality | | X | X | X | X | Respondent reported |
| | Vaccination history (DTP, Rotavirus, Polio, MMR) | | X | X | X | X | Observation of child health card |
| | Medication history | | X | X | X | X | Respondent reported |
| | Deworming history | | | | | X | Respondent reported |
| | Illness (diarrhea, dysentery, nasal congestion, fever, vomiting, physical injury) in the last week | | X | X | X | X | Respondent reported |
| | Medical diagnosis (colic, ear infection, anemia, respiratory illness, malaria, asthma) and treatment history | | X | X | X | X | Respondent reported |
| **Anthropometry** | Head circumference | | X | X | X | X | Objective measurement |
| | Body length | | X | X | X | X | Objective measurement |
| | Body weight | | X | X | X | X | Objective measurement |
| **Nutrition** | Types of liquid food consumed in the last week | | X | X | X | X | Respondent reported |
| | Types of solid food consumed in the last week | | | X | X | X | Respondent reported |
| **Sanitation** | Diaper wearing | | X | X | X | X | Respondent reported |
| | Disposal of child stools | | X | X | X | X | Respondent reported |
| **Water consumption** | Child consumption of drinking water source | | X | X | X | X | Respondent reported |
| | Drinking water treatment for child consumption | | X | X | X | X | Respondent reported |
| **Travel** | Estimated time spent away from home in the last 12 months | | | | | X | Respondent reported |

| Category | Indicator | Time point | | | | | Type of data |
|---|---|---|---|---|---|---|---|
| | | Pre-birth | 3mo | 6mo | 9mo | 12mo | |
| **Drinking water-level** | | | | | | | |
| **Water access** | Municipality water connection and status | X | X | X | X | X | Observation of water meter |
| | Type of drinking water source | X | X | X | X | X | Respondent reported<br>Observation of water source |
| | Alternate source for non-drinking water | X | X | X | X | X | Respondent reported |
| | Alternate source for drinking water outages | X | X | X | X | X | Respondent reported |
| | Distance to water main | X | X | X | X | X | GPS location |
| | Location of drinking water source | X | X | X | X | X | Respondent reported |
| | Distance to drinking water source | X | X | X | X | X | GPS location |
| | Time to collect water (minutes for round trip and trips per week) | X | X | X | X | X | Respondent reported |
| **Water availability** | Water availability (hours per day and days per week) | X | X | X | X | X | Respondent reported |
| | Household water storage (large and small containers) | X | X | X | X | X | Respondent reported |
| | Piped water flow rate (liters per minute) | X | X | X | X | X | Objective measurement |
| | Water availability at handwashing facility | X | X | X | X | X | Observation of handwashing facility |
| | Water availability at sanitation facility | X | X | X | X | X | Observation of sanitation facility |
| **Drinking water quality** | Treatment of stored water | X | X | X | X | X | Respondent reported |
| | Drinking water free/total chorine (source and stored water) | X | X | X | X | X | Objective measurement |
| **Water security** | Sharing of water connection | X | X | X | X | X | Respondent reported |
| | Household water insecurity experiences | X | X | X | X | X | Respondent reported |
| | Sufficient quantity of drinking water | X | X | X | X | X | Respondent reported |
| **User satisfaction with water** | Satisfied with water service | X | X | X | X | X | Respondent reported |
| | Satisfied with water availability | X | X | X | X | X | Respondent reported |
| | Satisfied with water pressure | X | X | X | X | X | Respondent reported |
| | Satisfied with water color and appearance | X | X | X | X | X | Respondent reported |
| | Satisfied with water taste and smell | X | X | X | X | X | Respondent reported |
| | Satisfied with water affordability | X | X | X | X | X | Respondent reported |
| **Water consumption** | Monthly water expenses | X | X | X | X | X | Respondent reported |
| | Water usage (liters per day) | X | X | X | X | X | Respondent reported<br>Observation of water meter |

### Table S2: Targets Assayed by the TaqMan Array Card (TAC) for the PAASIM study

| Target | Gene of interest | Forward (5' to 3') | Reverse (5' to 3') | Probe (5' to 3')* |
|---|---|---|---|---|
| | | **Bacterial Pathogens** | | |
| Enteroaggregative *E. coli* (EAEC) | aaiC | ATTGTCCTCAGGCATTTCAC | ACGACACCCCTGATAAACAA | TAGTGCATACTCATCATTTAAG |
| | aatA | CTGGCGAAAGACTGTATCAT | TTTTGCTTCATAAGCCGATAGA | TGGTTCTCATCTATTACAGACAGC |
| Diarrheagenic *E. coli* (DAEC) | afaB | GTCTCCCTGAATGTACAGCTTTCA | CMCTCTGCCACTCCACCTT | TCAAGCTGTTTGTTCGTC |
| Shiga toxin-producing *E. coli* (STEC) | stx1 | ACTTCTCGACTGCAAAGACGTATG | ACAAATTATCCCCTGWGCCACTATC | CTCTGCAATAGGTACTCCA |
| | stx2 | CCACATCGGTGTCTGTTATTAACC | GGTCAAAACGCGCCTGATAG | TTGCTGTGGATATACGAGG |
| Enteropathogenic *E. coli* (EPEC) | eae | CATTGATCAGGATTTTTCTGGTGATA | CTCATGCGGAAATAGCCGTTA | ATACTGGCGAGACTATTTCAA |
| | bfpA | TGGTGCTTGCGCTTGCT | CGTTGCGCTCATTACTTCTG | CAGTCTGCGTCTGATTCCAA |
| Enterotoxigenic *E. coli* (ETEC) | LT | TTCCCACCGGATCACCAA | CAACCTTGTGGTGCATGATGA | CTTGGAGAGAAGAACCCT |
| | STh | GCTAAACCAGCAGGGTCTTCAAAA | CCCGGTACAAGCAGGATTACAACA | TGGTCCTGAAAGCATGAA |
| | STp | TGAATCACTTGACTCTTCAAAA | GGCAGGATTACAACAAAGTT | TGAACAACACATTTTACTGCT |
| *Shigella* spp./ Enteroinvasive *E. coli (EIEC)* | ipaH | CCTTTTCCGCGTTCCTTGA | CGGAATCCGGAGGTATTGC | CGCCTTTCCGATACCGTCTCTGCA |
| *E. coli* O157 | rbdE | TTTCACACTTATTGGATGGTCTCAA | CGATGAGTTTATCTGCAAGGTGAT | CTCTCTTTCCTCTGCGGTCCT |
| *Campylobacter jejuni/ C. coli* | cadF | CTGCTAAACCATAGAAATAAAATTTCTCAC | CTTTGAAGGTAATTTAGATATGGATAATCG | CATTTTGACGATTTTTGGCTTGA |
| *Salmonella* spp. | ttr | CTCACCAGGAGATTACAACATGG | AGCTCAGACCAAAAGTGACCATC | CACCGACGGCGAGACCGACTTT |
| *Vibrio cholerae* | hylA | ATCGTCAGTTTGGAGCCAGT | TCGATGCGTTAAACACGAAG | ACCGATGCGATTGCCCAA |
| *Clostridium difficile* | tcdB | GGTATTACCTAATGCTCCAAATAG | TTTGTGCCATCATTTTCTAAGC | CCTGGTGTCCATCCTGTTTC |
| | | **Viral Pathogens** | | |
| Adenovirus | fiber | AACTTTCTCTCTTAATAGACGCC | AGGGGGCTAGAAAACAAAA | CTGACACGGGCACTCT |
| Astrovirus | capsid | CAGTTGCTTGCTGCGTTCA | CTTGCTAGCCATCACACTTCT | CACAGAAGAGCAACTCCATCGC |
| Hepatitis G | 5’ UTR | CGGCCAAAAGGTGGT GGA TG | CGACGAGCCTGACGTCGGG | AGGTCCCTCTGGCGCTTGTGGCGAG |
| Norovirus GI | ORF1-ORF2 | CGCTGGATGCGATTCCATGA | CTTAGACGCCATCATCATTTAC | TGGACAGGAGATCGC |
| Norovirus GII | ORF1-ORF2 | CAAGAACCTATGTTTAGATGGATGAG | TCGACGCCATCTTCATTCACA | TGGGAGGGCGATCGCAATCT |
| Rotavirus | NSP3 | ACCATCTWCACRTRACCCTCTATGAG | GGTCACATAACGCCCCTATAGC | AGTTAAAAGCTAACACTGTCAAA |
| Sapovirus | RdRp | GAYCASGCTCTCGCYACCTAC | CCCTCCATYTCAAACACTA; TTGGCCCTCGCCACCTAC | CCRCCTATRAACCA |
| SARS-CoV-2 | N1 | GACCCCAAAATCAGCGAAAT | TCTGGTTACTGCCAGTTGAATCTG | ACCCCGCATTACGTTTGGTGGACC |

| Target | Gene of interest | Forward (5' to 3') | Reverse (5' to 3') | Probe (5' to 3')* |
|---|---|---|---|---|
| | | **Protozoan pathogens** | | |
| *Cryptosporidium spp.* | 18S rRNA | GGGTTGTATTTATTAGATAAAGAACCA | AGGCCAATACCCTACCGTCT | TGACATATCATTCAAGTTTCTGAC |
| *Giardia duodenalis* | 18S rRNA | GACGGCTCAGGACAACGGTT | TTGCCAGCGGTGTCCG | CCCGCGGCGGTCCCTGCTAG |
| *Entamoeba histolytica* | 18S rRNA | ATTGTCGTGGCATCCTAACTCA | GCGGACGGCTCATTATAACA | TCATTGAATGAATTGGCCATTT |
| *Cyclospora cayetanensis* | 18S rRNA | AAAAGCTCGTAGTTGGATTTCTG | AACACCAACGCACGCAGC | AAGGCCGGATGACCACGA |
| | | **Helminthic pathogens** | | |
| *Ascaris lumbricoides* | ITS1 | GCCACATAGTAAATTGCACACAAAT | GCCTTTCTAACAAGCCCAACAT | TTGGCGGACAATTGCATGCGAT |
| *Trichuris trichiura* | 18S rRNA | TTGAAACGACTTGCTCATCAACTT | CTGATTCTCCGTTAACCGTTGTC | CGATGGTACGCTACGTGCTTACCATGG |
| *Ancylostoma duodenale* | ITS2 | GAATGACAGCAAACTCGTTGTTG | ATACTAGCCACTGCCGAAACGT | ATCGTTTACCGACTTTAG |
| *Necator americanus* | ITS2 | CTGTTTGTCGAACGGTACTTGC | ATAACAGCGTGCACATGTTGC | CTGTACTACGCATTGTATAC |
| | | **Antimicrobial resistance genes** | | |
| intl1 | | GATCGGTCGAATGCGTGT | GCCTTGATGTTACCCGAGAG | ATTCCTGGCCGTGGTTCTGGGTTTT |
| mcr-1 | | GATCGCTGTCGTGCTCTTTG | ACCGCGCCCATGATTAATAG | CGATGCTACTGATCACCACG |
| SHV | | TCCCATGATGAGCACCTTTAAA | TCCTGCTGGCGATAGTGGAT | TGCCGGTGACGAACAGCTGGAG |
| TEM | | GCATCTTACGGATGGCATGA | GTCCTCCGATCGTTGTCAGAA | CAGTGCTGCCATAACCATGAGTGA |
| CTX-M1 | | CCGTCACGCTGTTRTTAGGA | AATGCCACMCCCAGYCKKCC | CAGCAAAAACTTGCCGRATT |
| CTX-M8-M25 | | ATRACACSTTCCGGCTCGAT | GCTAAYGGCGTGGTGGTATC | TCAACACCGCGATCCCCG |
| CTX-M2-M74 | | GCGCAGACCCTGAAAAAYCT | TGYGCSCGCTGRGTTTCC | ACSCTGGGYAAAGCGC |
| CTX-M9 | | GCTTTATGCGCAGACGARTG | ATCACCGCGATAAAGCACCT | TCGATACCRMAGATAATACGC |
| KPC | | GGCCGCCGTGCAATAC | GCCGCCCAACTCCTTCA | TGATAACGCCGCCGCCAATTTGT |
| NDM | | ATATCACCGTTGGGATCGAC | TAGTGCTCAGTGTCGGCATC | AAGGACAGCAAGGCCAAGTCG |
| VIM | | TSTACCCRTCCAATGGTCTC | AGAAGKGCCRCTGTGTTTTT | TGTCCGTGATGGYGATGAGTTG |
| | | **Controls** | | |
| 16S | | TGCAAGTCGAACGAAGCACTTTA | GCAGGTTACCCACGCGTTAC | CGCCACTCAGTCACAAA |
| human mtDNA | | CAATGAATCTGAGGAGGCTAC | CGTGCAAGAATAGGAGGTG | ACCCTCACACGATTCTTTACCTTTCACT |
| BHV | | GAGCAAAGCCCCGCCGAAGGA | TACGAACAGCAGCACGGGCGG | GAACCTGCCCACGCGCTGAAAC |
| BRSV | | GCAATGCTGCAGGACTAGGTATAAT | ACACTGTAATTGATGACCCCATTCT | ACCAAGACTTGTATGATGCTGCCAAAGCA |

*All probes have FAM on the 5’ end and MGB on the 3’ end

**Table S3: Details of Power Calculations**

The table below details our minimum detectible effect for individual and groups of pathogens given the control group prevalence (p1) ranging from 10% to 80%. It generates the minimum detectible effect as a risk ratio (delta) and resulting minimum detectible prevalence in the comparison group.

Our assumptions are the number of sub-neighborhoods in the intervention and comparison (M1/M2), average number of households enrolled per sub-neighborhood in intervention and control (K1/K2), standard alpha (0.05) and power (.80). We estimate the minimum detectible effect for three values of for the intra-class correlation coefficient, representing low, moderate and high clustering ($\rho$ = .01, 0.05, 0.1). Our power calculations rely on the moderate estimates for ICC. We account for clustering at the sub-neighborhood level.

We apply the following code using STATA v16:

*power twoproportions (.2(.05).8) , cluster effect(ratio) m1(36) m2(26) k1(8) k2(10) power(.8) alpha(.05) direction(lower) rho(.01/.5/.1)*

| alpha | power | K1 | K2 | M1 | M2 | delta | p1 | p2 | rho |
|---|---|---|---|---|---|---|---|---|---|
| .05 | .8 | 8 | 10 | 36 | 26 | .5120 | .2 | .1024 | .01 |
| .05 | .8 | 8 | 10 | 36 | 26 | .5676 | .25 | .1419 | .01 |
| .05 | .8 | 8 | 10 | 36 | 26 | .6114 | .3 | .1834 | .01 |
| .05 | .8 | 8 | 10 | 36 | 26 | .6476 | .35 | .2266 | .01 |
| .05 | .8 | 8 | 10 | 36 | 26 | .6785 | .4 | .2714 | .01 |
| .05 | .8 | 8 | 10 | 36 | 26 | .7058 | .45 | .3176 | .01 |
| .05 | .8 | 8 | 10 | 36 | 26 | .7302 | .5 | .3651 | .01 |
| .05 | .8 | 8 | 10 | 36 | 26 | .7527 | .55 | .414 | .01 |
| .05 | .8 | 8 | 10 | 36 | 26 | .7736 | .6 | .4642 | .01 |
| .05 | .8 | 8 | 10 | 36 | 26 | .7935 | .65 | .5158 | .01 |
| .05 | .8 | 8 | 10 | 36 | 26 | .8127 | .7 | .5689 | .01 |
| .05 | .8 | 8 | 10 | 36 | 26 | .8317 | .75 | .6238 | .01 |
| .05 | .8 | 8 | 10 | 36 | 26 | .8509 | .8 | .6807 | .01 |
| .05 | .8 | 8 | 10 | 36 | 26 | .3585 | .2 | .07169 | .05 |
| .05 | .8 | 8 | 10 | 36 | 26 | .4262 | .25 | .1066 | .05 |
| .05 | .8 | 8 | 10 | 36 | 26 | .4804 | .3 | .1441 | .05 |
| .05 | .8 | 8 | 10 | 36 | 26 | .5257 | .35 | .184 | .05 |
| .05 | .8 | 8 | 10 | 36 | 26 | .5648 | .4 | .2259 | .05 |
| .05 | .8 | 8 | 10 | 36 | 26 | .5994 | .45 | .2698 | .05 |
| .05 | .8 | 8 | 10 | 36 | 26 | .6308 | .5 | .3154 | .05 |
| .05 | .8 | 8 | 10 | 36 | 26 | .6596 | .55 | .3628 | .05 |
| .05 | .8 | 8 | 10 | 36 | 26 | .6867 | .6 | .412 | .05 |
| .05 | .8 | 8 | 10 | 36 | 26 | .7126 | .65 | .4632 | .05 |
| .05 | .8 | 8 | 10 | 36 | 26 | .7376 | .7 | .5163 | .05 |
| .05 | .8 | 8 | 10 | 36 | 26 | .7624 | .75 | .5718 | .05 |
| .05 | .8 | 8 | 10 | 36 | 26 | .7875 | .8 | .63 | .05 |
| .05 | .8 | 8 | 10 | 36 | 26 | .5398 | .5 | .2699 | .1 |
| .05 | .8 | 8 | 10 | 36 | 26 | .5737 | .55 | .3155 | .1 |
| .05 | .8 | 8 | 10 | 36 | 26 | .6056 | .6 | .3634 | .1 |
| .05 | .8 | 8 | 10 | 36 | 26 | .6362 | .65 | .4135 | .1 |
| .05 | .8 | 8 | 10 | 36 | 26 | .6660 | .7 | .4662 | .1 |
| .05 | .8 | 8 | 10 | 36 | 26 | .6955 | .75 | .5216 | .1 |
| .05 | .8 | 8 | 10 | 36 | 26 | .7254 | .8 | .5804 | .1 |
| .05 | .8 | 8 | 10 | 36 | 26 | .7567 | .85 | .6432 | .1 |
| .05 | .8 | 8 | 10 | 36 | 26 | .7907 | .9 | .7117 | .1 |

10. De Onis M. WHO Child Growth Standards: Length/height-for-age, weight-for-age, weight-for-length, weight-for-height and body mass index-for-age: Methods and development. Geneva, Switzerland: World Health Organization; 2006.

11. World Health Organization. WHO child growth standards, SAS macros. Geneva, Switzerland: World Health Organization; 2010.

20. Centers for Disease Control and Prevention Division of Parasitic Diseases and Malaria. Blood Collection and Dried Blood Spot Preparation. Atlanta, GA: Centers for Disease Control and Prevention; 2019. Report No.: ECL.DR.C.032.

21. Lammie PJ, Moss DM, Brook Goodhew E, Hamlin K, Krolewiecki A, West SK, et al. Development of a new platform for neglected tropical disease surveillance. Int J Parasitol. 2012 Aug;42(9):797–800.

22. Zambrano LD, Priest JW, Ivan E, Rusine J, Nagel C, Kirby M, et al. Use of Serologic Responses against Enteropathogens to Assess the Impact of a Point-of-Use Water Filter: A Randomized Controlled Trial in Western Province, Rwanda. Am J Trop Med Hyg. 2017 Sep;97(3):876–87.

23. Zinkernagel RM. Maternal antibodies, childhood infections, and autoimmune diseases. N Engl J Med. 2001 Nov;345(18):1331–5.

24. 4500-Cl CHLORINE (RESIDUAL). In: Standard Methods For the Examination of Water and Wastewater [Internet]. American Public Health Association; 2018 [cited 2021 Oct 11]. (Standard Methods for the Examination of Water and Wastewater). Available from: https://www.standardmethods.org/doi/abs/10.2105/SMWW.2882.078
